# Adherence to 24h movement recommendations and health indicators in the ABCD study: Cross-sectional and longitudinal associations with cognition, psychosocial health, BMI and brain structure

**DOI:** 10.1101/2022.03.23.22272692

**Authors:** Hoki Fung, B.T. Thomas Yeo, Christina Chen, June C. Lo, Michael W.L. Chee, Ju Lynn Ong

## Abstract

Adherence to 24h movement guidelines of ≥60min of physical activity, ≤2h of screen time, and 9-11h of sleep has been shown to benefit cognition, physical and psychosocial health in children aged 5-13y. However, these findings are largely based on cross-sectional studies or small samples. Here we utilize data from the Adolescent Brain Cognitive Development (ABCD) study of 10000+ children aged 9-11y to examine whether adherence to 24h movement guidelines benefit cognition, BMI, psychosocial health and brain morphometric measures at baseline (T1) and 2 years later (T2). After adjustment for sociodemographic confounders in multivariable linear mixed models, we observed better cognitive scores, fewer behavioural problems, lower adiposity levels and greater gray matter volumes in children who met both sleep and screen time recommendations compared to those who met none. Longitudinal follow up further supports these findings; participants who met both recommendations at T1 and T2 evidenced better outcome measures than those who met none, even after controlling for T1 measures. These findings support consideration of integrated rather than isolated movement recommendations across the day for better cognitive, physical, psychosocial and brain health.

## INTRODUCTION

Adequate sleep, regular physical activity, and limitation of sedentary behaviour (including screen time) are well-recognized determinants of healthy development in childhood and adolescence.^1-3^ While often dealt with separately, these modifiable lifestyle factors lie on a continuum of interacting ‘movement’ behaviours.^4^ For example, health benefits of regular levels of moderate-to-vigorous physical activity (MVPA) could be attenuated if a child engages in excessive screen time / sedentary behaviours and has inadequate sleep.^5^

Recognizing this, the WHO^6^ and several countries^4,7-9^ are shifting away from separate recommendations and moving towards the adoption of 24h movement guidelines for overall health and wellbeing. Based on a set of systematic reviews^10-12^ and guidelines first proposed by the Canadian task-force in 2016 (i.e. the Canadian 24-Hour Movement Guidelines for Children and Youth),^4^ 24h guidelines typically recommend at least 60min of MVPA per day, 2h or less recreational screen time per day, and 9-11h sleep per night for children aged 5-13y.

In support of these guidelines, recent studies have found positive associations between the number of recommendations met and overall health indicators in children and adolescents.^13^ Participants who fulfilled more guideline components had lower adiposity levels,^14,15^ higher health-related quality of life,^16-18^ better psychosocial^19^ and mental health scores,^20,21^ lower risk of depression,^22^ and superior academic performance^23^ as well as global cognition scores.^24^ However, these findings are mostly derived from cross-sectional studies or relatively small samples. Results have been equivocal, limited health indicators have been studied at a time, and the relationships between adherence to 24h movement guidelines and brain structure during this critical developmental period have not been studied. To answer these questions, we utilized data from a large sample of 10,000+ children aged 9-11y from the Adolescent Brain and Cognitive Development (ABCD) study, to investigate associations between adherence to movement guidelines from 24h movement behaviour data (24h-MB) surveying sleep, screen time and physical activity behaviours, and four commonly surveyed health indicators measuring: 1) cognition; 2) psychosocial health; 3) body mass index (BMI) and 4) brain morphometric measures, both at baseline and 2 years later.

## MATERIALS AND METHODS

### Data Source

The ABCD Study (http://abcdstudy.org) is the largest longitudinal study on brain development and child health in the United States. It involves 22 nationally distributed research sites and aims to follow 11,878 children from pre-adolescence into early adulthood. Potential participants were identified through probability sampling of public and private elementary schools estimated to cover over 20% of eligible participants in the U.S. population. Additional procedures such as summer recruitment and referrals were adopted to recruit harder-to-reach populations such as home-schooled children to achieve a near-representative and diverse national sample. Informed consent, assent, and parental permission were obtained prior to all data collection. Further details on recruitment procedures, task design, data collection protocols, and neuroimage processing pipelines are described elsewhere.^25,26^

Analyses were conducted on data from the fourth annual release (http://dx.doi.org/10.15154/1523041). After excluding participants who did not provide sociodemographic information for age, sex, highest parental education, household income and race, 10574 and 9273 participants contributed data on 24h movement behaviours (24h-MB) for the baseline (T1) and 2-year follow up (T2) timepoints respectively (Table 1). For each of the outcome variables considered, a further number of participants had to be excluded due to missing data and quality checks (Supplementary Table 1). As only de-identified data were used, this study was exempted from a full review by the Institutional Review Board at the National University of Singapore.

**Table 1.** Demographic, Behavioural, and Brain Volumetric Characteristics

|  | **T1 (9-11y)** | **T2 (11-14y)** |
| --- | --- | --- |
|  | **Total N = 10574** | **Total N = 9273** |
| **Demographics** | **N (%) or Mean (SD)** | **N (%) or Mean (SD)** |
| Sex |  |  |
| Male | 5504 (52.1%) | 4849 (52.2%) |
| Female | 5070 (48.0%) | 4424 (47.7%) |
| Age (y) | 9.9 (0.6) | 12.0 (0.7) |
| Highest parental education † | 16.8 (2.6) | 16.9 (2.5) |
| Family income ‡ | 7.3 (2.4) | 7.4 (2.3) |
| Race |  |  |
| African American | 1568 (14.8%) | 1259 (13.6%) |
| American Indian and Alaska Native | 54 (0.5%) | 44 (0.05%) |
| Asian | 234 (2.2%) | 197 (2.1%) |
| Pacific Islander | 13 (0.1%) | 11 (0.1%) |
| White | 6979 (66.0%) | 6267 (67.6%) |
| Other | 427 (4.0%) | 365 (3.9%) |
| Two or more races | 1299 (12.3%) | 1130 (12.1%) |
| **24h Movement Behaviours** |  |  |
| Sleep Duration |  |  |
| 9 – 11 h | 5166 (48.8%) | 3012 (32.5%) |
| 8 – 9 h | 3866 (36.6%) | 4085 (44.0%) |
| 7 – 8 h | 1211 (11.5%) | 1664 (17.9%) |
| 5 – 7 h | 312 (3.0%) | 474 (5.1%) |
| < 5 h | 29 (0.2%) | 38 (0.4%) |
| Weighted screen time (hours/day) | 3.7 (3.0) | 4.0 (3.2) |
| Moderate-to-vigorous physical activity (days/week) | 3.5 (2.3) | 3.8 (2.1) |
| **Cognition (NIH-Toolbox)** |  |  |
| Fluid Intelligence | 91.9 (10.6) | 93.7 (12.4) |
| Crystallised Intelligence | 86.7 (7.0) | 91.2 (7.1) |
| Mean Cognitive Score (5 Tasks) | 92.3 (6.5) | 99.1 (6.9) |
| **Psychosocial Health (Child Behaviour Checklist)** |  |  |
| Externalizing Problems | 45.7 (10.3) | 44.5 (9.7) |
| Internalizing Problems | 48.5 (10.6) | 47.8 (10.4) |
| Total Problems | 45.9 (11.2) | 44.8 (11.2) |
| **Body Mass Index (kg/m<sup>2</sup>)** | **18.7 (3.9)** | **20.3 (4.3)** |
| **Brain Volumes (mm<sup>3</sup>)** |  |  |
| Cortical Gray Matter Volume | 598740.3 (56117.6) | 590379.1 (56724.2) |
| Subcortical Gray Matter Volume | 60052.5 (4874.2) | 60861.6 (4945.8) |
| Intracranial Volume | 1494839 (143524.6) | 1525469 (147482.5) |

Highest parental education and family income were measured in ordinal scales. †Highest parental education referred to the highest level of school the parent had completed, 1-12=Grade 1 to 12; 13=High school graduate; 14=GED or equivalent; 15=Some college; 16=Associate degree: Occupational; 17=Associate degree: Academic; 18=Bachelor's degree; 19=Master's degree; 20=Professional School degree; and 21=Doctoral degree. ‡Family income referred to the total combined family income for the past 12 months before taxes and deductions, 1=Less than \$5,000; 2=\$5,000 through \$11,999; 3=\$12,000 through \$15,999; 4=\$16,000 through \$24,999; 5=\$25,000 through \$34,999; 6=\$35,000 through \$49,999; 7=\$50,000 through \$74,999; 8= \$75,000 through \$99,999; 9=\$100,000 through \$199,999; and 10=\$200,000 and greater.

### 24h Movement Behaviours (24h-MB)

Self and parental reports were used to extract the amount of time participants spent on each of the three movement behaviours (i.e. sleep, screen time and physical activity). These responses were then recoded as binary variables, with 1 indicating adherence, and 0 indicating non-adherence, to the 24h movement recommendations. Participants were subsequently grouped into 8 categories of adherence to each combination of 24h-MBs to examine specificity of these MB categories to the different outcome variables investigated (Figure 1).

**Figure 1.**
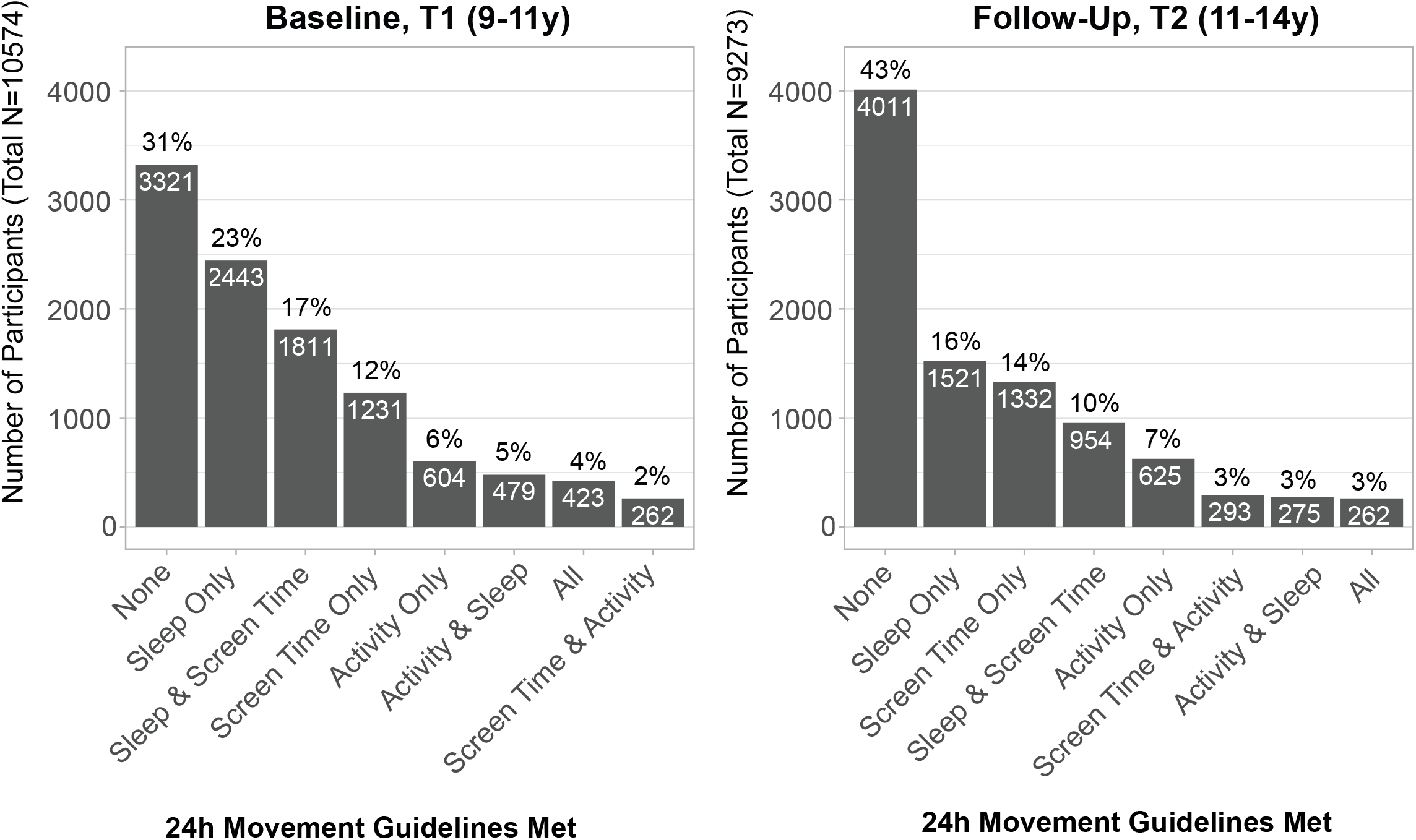
Number of children meeting 24h movement guidelines at baseline (T1) and follow up (T2).

Sleep duration was obtained from parental responses to an item in the Parent Sleep Disturbance Scale for Children^27^ that queried, “How many hours of sleep does your child get on most nights?”. Parents selected one of five available options: 9-11h; 8-9h; 7-8h; 5-7h; and <5h. Children who received 9-11h of sleep were deemed to have met the 24h movement recommendations.

Recreational screen time (a major contributor to sedentary behaviour^12^) was assessed using the Youth Screen Time Survey.^28^ At T1, participants were asked to report the number of hours spent in six categories: (1) TV shows or movies, (2) Videos (YouTube etc.), (3) Video games on a computer, console, phone or other device (Xbox, Play Station, iPad), (4) Text on a cell phone, tablet, or computer (GChat, WhatsApp, etc.), (5) Social networking sites (Facebook, Twitter, Instagram, etc.) and (6) Video chat (Skype, FaceTime, etc.) on a typical weekday and weekend day. Possible categorical responses were: none; <30min (coded as 15min); 30min; 1h; 2h; 3h; _≥_4h (coded as 4h). A weighted mean daily screen time was computed from the 12 values provided ([(total screen time on a weekday*5 + total screen time on a weekend*2)/7]). At T2, participants were asked to report total time spent on weekdays and weekends in these six categories combined, and a corresponding weighted value was computed using the 2 values provided ([(total screen time on a weekday*5 + total screen time on a weekend*2)/7]). Children who spent _≤_2h on screens were deemed to have met screen time recommendations.

Time spent on physical activity was derived from the children’s response to an item in the Youth Risk Behaviour Survey^29^ that asked, “During the past 7 days, on how many days were you physically active for a total of at least 60 minutes per day? (Add up all the time you spent in any kind of physical activity that increased your heart rate and made you breathe hard some of the time)”. Responses could vary between zero to seven days, and only those who indicated that they were active 7 days/week were considered to have met the activity recommendations.

### Cognitive measures

Cognition was assessed with the NIH Toolbox Cognition Battery that comprised seven tasks spanning six cognitive domains: attention, episodic memory, working memory, language, executive function, and processing speed.^30^ A crystallized intelligence composite score was computed from the Picture Vocabulary and Oral Reading tasks while a fluid intelligence composite score was computed from the other five tasks (Pattern Comparison, List Sorting Working Memory, Picture Sequence, Flanker and Dimensional Cart Sort). As two of these tasks – the Dimensional Cart Sort and List Sorting Working Memory were only conducted at T1, a composite fluid intelligence score could not be computed at T2. The remaining five tasks (Picture Vocabulary, Flanker, Picture Sequence, Pattern Comparison and Oral reading) were instead averaged together for a composite cognition score that could be compared across both timepoints. Uncorrected standard scores (M = 100, SD = 15) were used, which compare the score of the test taker to those in the normative sample (age 3 - 85y).

### Psychosocial health measures

Parental reports of problem behaviours over the past 6 months were collected using the Child Behaviour Checklist (CBCL) for ages 6-18y.^31,32^ Composite scores summarizing internalizing problems (those directed towards ‘self’, e.g. from anxiety, depression and social withdrawal scales), externalizing problems (those directed towards ‘others’, e.g. from rule-breaking and aggressive behaviour scales) as well as an overall composite for total problems were extracted. Raw scores were converted to norm-referenced T-scores (M = 50, SD = 10), with higher scores indicating greater levels of emotional and behavioural problems. Clinically significant elevations are typically denoted by T-scores _≥_ 65.

### Body Mass Index (BMI)

Participants’ height and weight were taken three times and averaged to obtain BMI scores using the formula weight (kg) / height (m^2^). Extreme values of < 11 or > 36 kg/m2 were excluded as possible data entry errors/outliers (∼1% of data).

### Brain morphometric measures

Scanning parameters, pre-processing and analytical pipelines are described in separate papers.^26,33^ Structural brain images were processed using FreeSurfer 7.1.1 (https://surfer.nmr.mgh.harvard.edu) to obtain measures of estimated total intracranial volume (ICV), total cortical and subcortical gray matter volumes (GMVs). Only participants whose structural data passed quality review checks by the ABCD Study team (imgincl_t1w_include==1) were included in the final analysis.

### Statistical Analysis

Cross-sectional analyses were conducted for both T1 and T2 separately. Associations between adherence to each 24h-MB category (compared to a reference group meeting no recommendations) and outcome variables for cognition, psychosocial health, BMI and GMVs were investigated using multi-variable linear mixed effects models, controlled for age, sex, parental education, socioeconomic status and race. GMV models additionally controlled for ICV measures. As the ABCD study includes siblings, families nested within site were additionally modelled as a random effect. Unstandardized regression coefficients (b) are reported throughout the manuscript to show the difference between the mean of outcomes measured in each 24h-MB group to the mean of the reference group who did not meet any recommendations, keeping all other variables constant.

Next, longitudinal analyses were conducted to look at associations between changes from T1 to T2 (i.e. T2-T1) for each of the outcome variables considered, controlled for sociodemographic variables at T1 included in the cross-sectional analyses, outcomes at T1, and time elapsed between T1 and T2 assessments. Analyses were conducted first by comparing groups who met specific 24h-MB categories at both T1 and T2 with the group who did not meet any recommendations. As adherence to both sleep and screen time recommendations at both T1 and T2 most consistently showed favorable outcomes compared to the group who met no recommendations, we followed up this finding by also comparing groups who only met the sleep and screen time combination at *either* T1 or T2, compared to the rest of the sample.

p-values were corrected for multiple comparisons using the Benjamini-Hochberg procedure to control the False Discovery Rate (FDR) at 0.05.^34^ This correction was carried out separately for T1 (9 outcomes x 7 24h-MB categories), T2 (8 outcomes x 7 24h-MB categories) and T2-T1 (8 outcomes x 7 24h-MB categories). Results are reported with and without this correction.

### Post-Hoc Analyses

To follow-up on significant associations between adherence to each 24h-MB category and cortical GMV, we further inspected relationships by lobar region (frontal, parietal, temporal, occipital) following delineations in https://surfer.nmr.mgh.harvard.edu/fswiki/CorticalParcellation. To follow-up on unexpected findings between adherence to physical activity recommendations and a) composite cognition scores, b) externalizing behaviours and c) cortical GMV, we modelled frequency of MVPA per week as a categorical variable, with 0 days per week as the reference value in both T1 and T2, and additionally controlled for whether sleep and screen time recommendations were met (0/1 indicator variable) in addition to the other sociodemographic covariates used in the previous analyses.

All analyses were conducted using R version 4.1.0.

## RESULTS

Sample characteristics of the 10574 children (M ± SD = 9.9 ± 0.6y, 52% male) and 9273 (M ± SD = 12.0 ± 0.7y, 52% male) who provided demographic and 24h-MB data at T1 and T2 respectively are provided in Table 1. At T1, almost half of the sample (49%) met sleep guidelines of 9-11h/night, 35% met screen time guidelines of _≤_ 2h/day, and only 17% met MVPA guidelines of _≥_ 60min/day. Only 4% of participants met all three 24-h movement recommendations, while of concern, 31% did not meet any (Figure 1). At T2, the proportion of those who met sleep guidelines dropped to about a third (32%) of the sample, while the proportion meeting screen time (31%) and MVPA (16%) guidelines remained relatively stable. Notably, of the 9255 participants who provided 24h-MB data both at T1 and T2, only <1% met all three guidelines both at T1 and T2, while 20% did not meet any.

### Cross-sectional analyses

#### 24h Movement Behaviours and Cognition

At T1, compared to participants who did not meet any recommendations, those who met screen time only, sleep and screen time, or all three recommendations evidenced superior fluid intelligence, crystallized intelligence and composite cognition scores, with adherence to sleep and screen time recommendations showing the greatest benefit (b = 1.93, p < 0.001, p_FDR-corr_ < 0.001; Figure 2A). At T2 however, these benefits were not significant in any group (p > 0.11). Unexpectedly, children who met activity only (b= −0.94, p = 0.001, p_FDR-corr_ = 0.006), sleep and activity (b = −1.20, p = 0.007, p_FDR-corr_ = 0.019) and screen time and activity (b = −0.97, p = 0.013, p_FDR-corr_ = 0.03) recommendations instead showed poorer crystallised intelligence scores compared to those who did not meet any recommendations.

**Figure 2.**
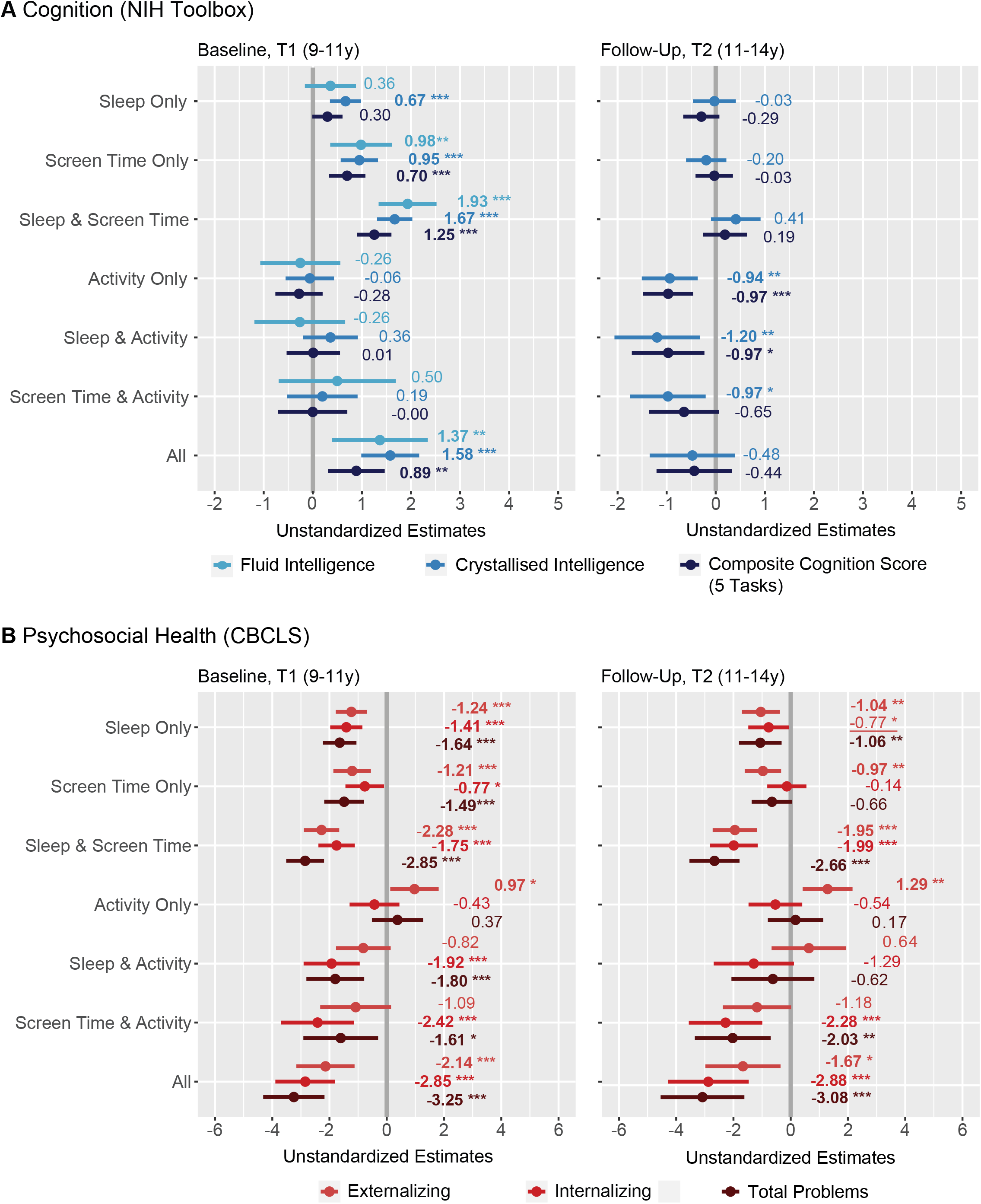
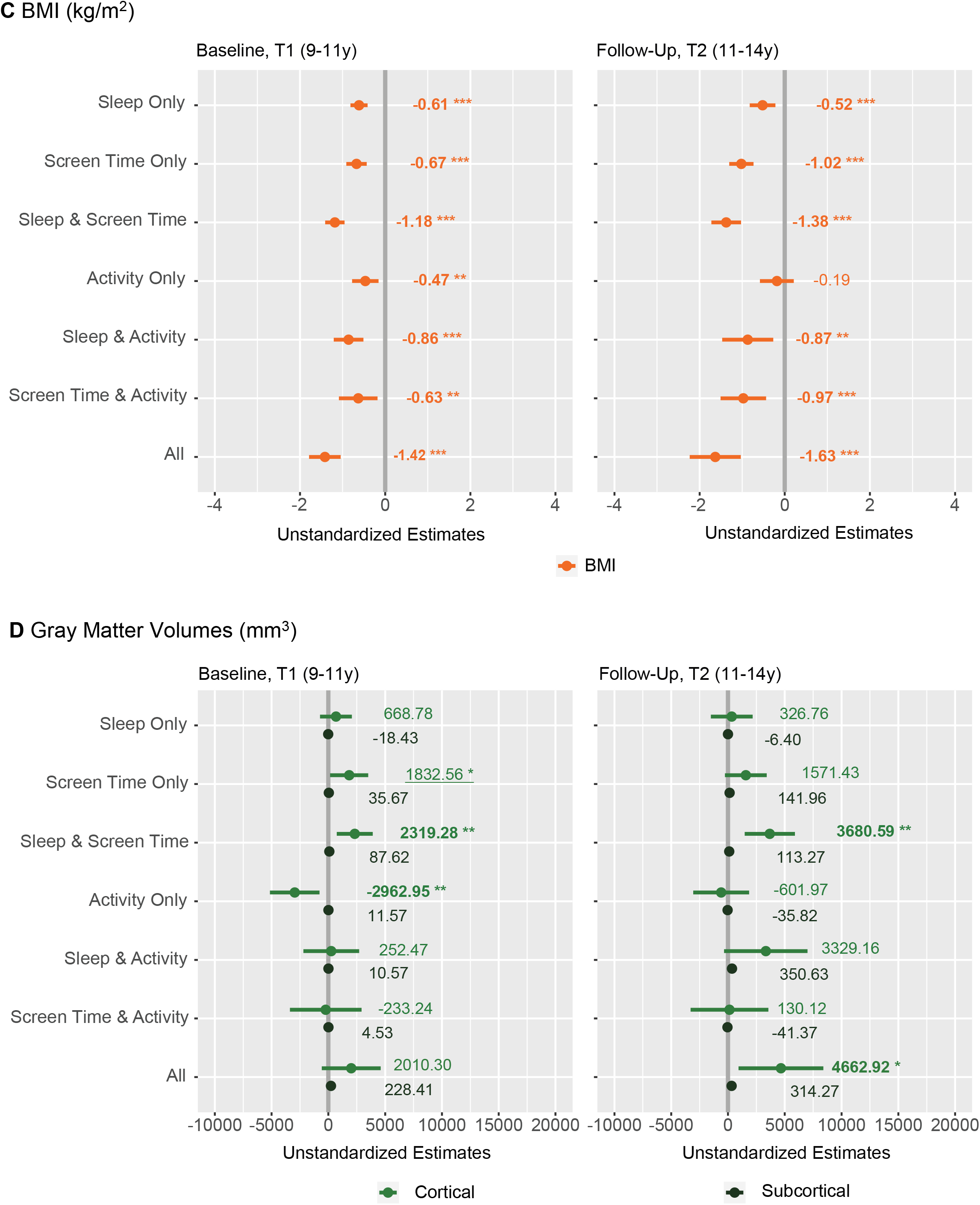
Cross-sectional associations between adherence to 24h movement behaviours and (A) cognitive scores, (B) psychosocial health scores, (C) BMI and (D) gray matter volumes (GMV). Plots show unstandardized estimates of regression coefficients from multilevel models for baseline (T1; left panel) and follow-up (T2; right panel) timepoints respectively. Error bars indicate 95% confidence intervals while asterisks denote level of significance, uncorrected (‘***’ p< 0.001 ‘**’ p < 0.01 ‘*’ p< 0.05). Significant p-values (p<0.05) that survive multiple comparisons corrections are denoted in bold font, while those that do not are underlined. The gray reference line refers to the group who did not meet any recommendations. All models were controlled for age, sex, highest level of education (parent), race, household income and intracranial volume (GMV models only). Families nested within site were additionally modelled as a random effect.

#### 24h Movement Behaviours and Psychosocial Health

At T1, participants who met most combinations of 24h-MBs evidenced lower internalizing and total problems compared to participants who did not meet any recommendations (Figure 2B). However, participants who met activity only recommendations evidenced greater levels of externalizing problems (b = 0.97, p = 0.02, p_FDR-corr_ = 0.04; Table 2). These associations were largely mirrored at T2, with the exception of those meeting sleep and activity and screen time only recommendations no longer evidencing lower levels of internalizing and total problems compared to the group who did not meet any recommendations.

#### 24h Movement Behaviours and BMI measures

At both T1 and T2, participants who met most combinations of 24h-MBs showed lower adiposity levels than those who did not meet any recommendations, with the exception of the group who only met activity recommendations at T2 (p = 0.36; Figure 2C).

#### 24h Movement Behaviours and Gray Matter Volumes

Compared to participants who did not meet any recommendations, only those who met both sleep and screen time recommendations alone consistently showed greater total cortical GMVs at both T1 (b = 2319.28 mm^3^, p = 0.004, p_FDR-corr_ = 0.009) and T2 (b = 3680.59 mm^3^, p=0.001, p_FDR-corr_ = 0.005) (Figure 2D). Follow-up analyses show that these relationships were significant in the frontal (T1: b = 876.68 mm^3^, p = 0.02; T2: b = 1449.00 mm^3^, p = 0.005) and temporal lobe regions (T1: b = 644.11 mm^3^, p = 0.004; T2: 903.47 mm^3^, p = 0.003) at both timepoints (Supplementary Figure 1). Those who only met activity recommendations at T1 evidenced smaller GMVs than those who did not meet any (b = −2962.95 mm^3^, p = 0.008, p_FDR-corr_ = 0.01) but this relationship was no longer significant at T2 (p = 0.63).

### Longitudinal Analyses

Considering groups of participants who met the same combinations of 24h-MBs at both T1 and T2 (Figure 3), it was clear that only the group who met sleep and screen time recommendations at both T1 and T2 showed improvements across time (T2-T1) on all health indicators surveyed. They showed the greatest increase in composite cognition score (b = 0.66, p = 0.02, p_FDR-corr_ = 0.21), decrease in externalizing (b = − 1.05, p = 0.02, p_FDR-corr_ = 0.18) and total problems (b = −1.23, p = 0.01, p_FDR-corr_ = 0.18), decrease in BMI (b = −0.60 kg/m^2^, p < 0.001, p_FDR-corr_ < 0.001) and increase in total cortical (b = 3242.37 mm^3^, p < 0.001, p_FDR-corr_ < 0.001) and subcortical (b =, cortical 139.04 mm^3^, p = 0.01, p_FDR-corr_ = 0.18) volumes compared to the group who did not meet any recommendations, although only BMI and total cortical change remained significant after FDR-correction. Follow-up analyses on groups who only met sleep and screen time recommendations at either T1 or T2 suggested that for BMI and GMV measures, early adherence (T1) to recommendations still benefitted T2-T1 measures (BMI: b = −0.17 kg/m^2^ p = 0.04; GMV: b = 1312.04 mm^3^, p = 0.001, b_subcortical_ = 100.15 mm^3^, p = 0.002) (Supplementary Figure 2). This suggests that some effects span a longer timescale. In contrast, for psychosocial health, T2 adherence to recommendations was related to improved T2-T1 outcomes (b_externalizing_ = −0.73, p = 0.04; b_internalizing_ = −0.89, p = 0.03; b_total_ = −0.94, p = 0.03), suggesting a more immediate effect. For cognition, only those who met sleep and screen time recommendations at both timepoints evidenced improvements (b = 0.5, p = 0.045) compared to the rest of the sample.

**Figure 3.**
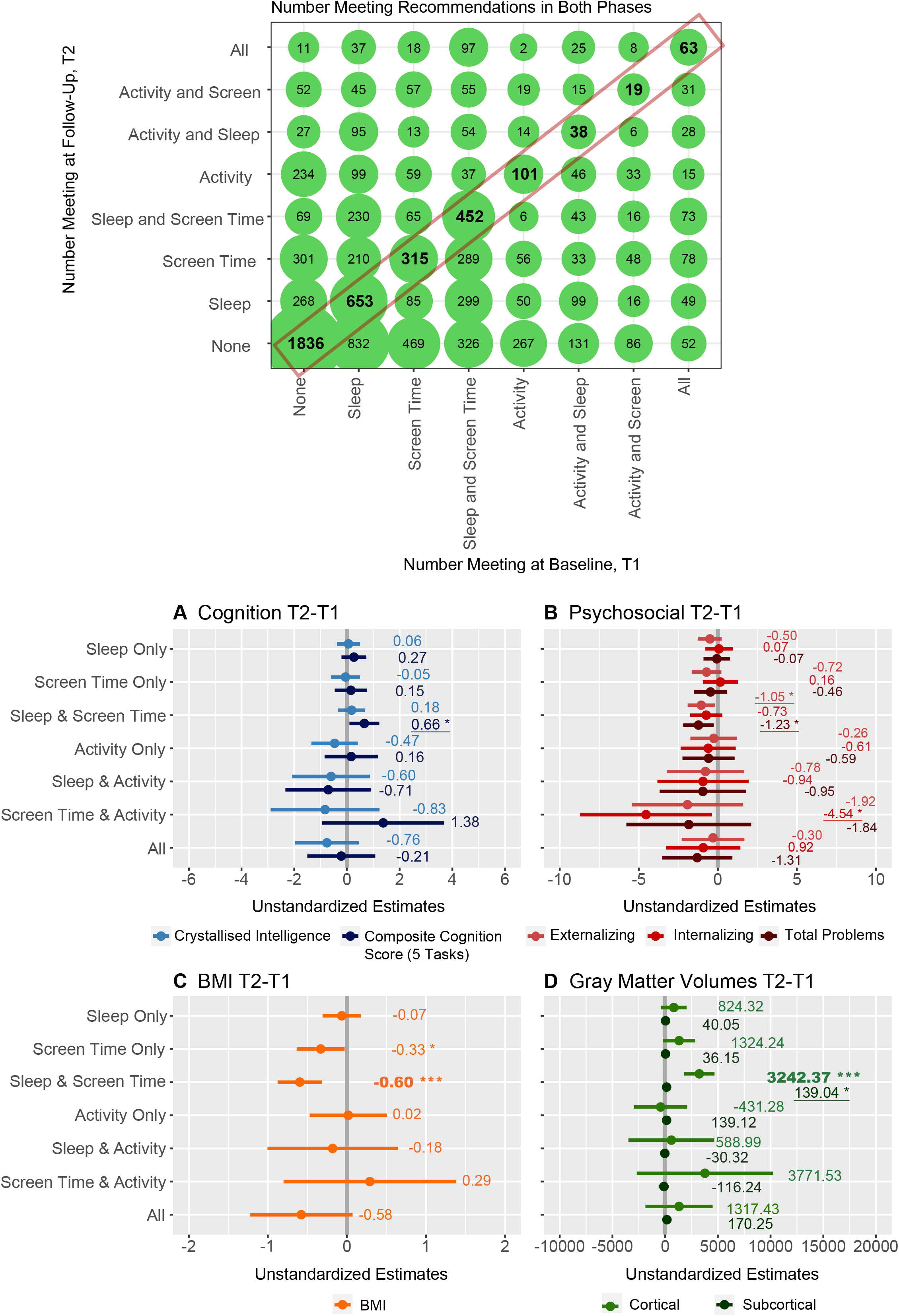
Top panel: Balloon plot showing frequencies of children meeting combinations of 24h recommendations at baseline (T1) and follow-up (T2). Bottom panel: Longitudinal (T2-T1) associations between adherence to 24h movement behaviours at both time points (i.e. groups indicated in the red rectangle) and (A) cognitive scores, (B) psychosocial health scores, (C) BMI and (D) gray matter volumes (GMV). Plots show unstandardized estimates of regression coefficients from multilevel models. The gray reference line refers to the group who did not meet any recommendations. Error bars indicate 95% confidence intervals while asterisks denote level of significance, uncorrected (‘***’ p< 0.001 ‘**’ p < 0.01 ‘*’ p< 0.05). Significant p-values (p<0.05) that survive multiple comparisons corrections are denoted in bold font, while those that do not are underlined. All models were controlled for age, sex, highest level of education (parent), race, household income, intracranial volume (GMV models only) as well as time elapsed between baseline and follow-up and baseline measures of all dependent variables. Families nested within site were additionally modelled as a random effect.

#### Post-Hoc Analyses

When frequency of physical activity per week was coded as an interval variable (0 – 7 days), non-linear relationships between frequency of physical activity per week and composite cognition, cortical GMV and externalizing behaviours were found (Supplementary Figure 3) whereby better health outcomes were associated with moderate levels of weekly physical activity.

## DISCUSSION

In this study, we found that children aged 9-11y who both slept 9-11h *and* kept to 2h or less recreational screen time a day demonstrated higher cognitive scores, better psychosocial health, lower adiposity levels and greater cortical GMVs than those who did not meet any recommendations. Approximately two years later, most of these associations persisted, although the association with cognition was not significant. Considering longitudinal changes, only those who met sleep and screen time recommendations at both timepoints again evidenced improvements across time on all health indicators than the group who did not meet any recommendations, even after controlling for baseline metrics, although only effects on BMI and gray matter volumes survived comparisons for multiple corrections.

While meeting either sleep or screen time recommendations alone was also beneficial for certain aspects of cognition, the effect was the largest when both recommendations were met. Sleep and screen time are closely linked, and an increase in screen time particularly before bedtime is known to negatively impact subsequent sleep.^35^ This is effected by delaying bedtimes, increasing sleep onset latency, awakenings after sleep onset and sleep disturbance severity, and reducing total sleep time,^35-37^ although we did not have information about timing of screen use in this dataset to investigate this further. Proposed mechanisms include displacement of time that would have been spent sleeping, delayed melatonin secretion due to evening light exposure, and increased pre-sleep arousal levels.^38^

Considering psychosocial health scores, children who met combinations including sleep and screen time also tended to show fewer internalizing, externalizing and total problems scores compared to those who did not. Adequate sleep has been shown to be important for emotional regulation and mental wellbeing and has been linked to fewer behavioural problems in school-aged children,^10,39^ while high levels of recreational screen time (>2h/day) have been associated with greater externalizing problems and psychological distress which could be linked to the content of screen use.^39^

For adiposity levels measured by body mass index scores, most combinations of adherence to recommendations (with the exception of activity at T2) resulted in lower BMI levels compared to those who met none, with the group who met all 24h-movement recommendations showing the lowest BMI on average compared to the group who met no recommendations. Sufficient sleep, together with a proper diet and adequate levels of physical activity are known to regulate hormones related to growth, maturation and energy homeostasis, while insufficient levels could lead to energy imbalance and obesity^40^ As higher BMI has been associated with poorer cognitive function across the lifespan^41^ as well as a range of long-term health issues,^42^ BMI levels need to be monitored throughout childhood and adolescence, and intervened appropriately.

Brain structural findings revealed that it was again the combination of sleep and screen time recommendations which consistently yielded greater GMVs compared to the group who met none, both cross-sectionally and longitudinally. Gray matter densities typically increase from birth before reaching a peak at around the onset of puberty, before declining throughout adolescence and early adulthood,^43-45^ suggestive of pruning or reorganization for efficient neural processing. This could suggest premature GMV decreases in children who do not meet/consistently meet optimal movement recommendations. Post-hoc analyses show that these cross-sectional differences tended to occur in the frontal and temporal regions - areas that mature later and could be most vulnerable to insults during this period, while longitudinal differences were more widespread across the brain. While prior neuroimaging work has focused separately on associations between 24h-MB components (e.g. sleep^20^ or screen time^46,47^) and brain structure, these findings suggest that these movement behaviours need to be simultaneously targeted for better developmental outcomes.

Intriguingly, combinations with adherence to activity recommendations appeared to have either non-significant or even detrimental effects on some of the health indicators surveyed here (unless they were combined with adherence to both sleep and screen time recommendations). For cognition and GMV, this could seem contradictory to many cross-sectional studies which show a positive association between physical fitness and academic success,^48^ likely through a facilitation of neuroplasticity on certain brain structures and consequently cognitive function.^49^ Considering that this group also exhibited more externalizing problems at both T1 and T2, this could suggest a coping mechanism in children with behavioural disorders who are also more likely to be hyperactive, act out more and may also be more attracted to physical activity, or that their parents themselves encourage physical activity as a means to dealing with behavioural issues.^20^ Children with higher symptom attention-deficit/hyperactivity-disorder severity have been shown to exhibit poorer cognition and smaller GMV in widespread regions.^50^ In addition, post-hoc analyses at T1 showed a beneficial relationship between meeting physical activity recommendations for moderate levels of physical activity (e.g. 1-6 days/week compared to 0 days/week but not at 7 days for cognition) suggesting that this benefit could have been accruing with lesser days of adherence. This finding supports recent guideline updates which explicitly state that children can accumulate physical activity through “an average of 60 min per day of MVPA per day and not necessarily on all 7 days of the week”.^51^

Our study clearly highlights the contributions of 24h movement behaviours to cognitive, mental and physical health in a very large sample and from both cross-sectional and longitudinal perspectives. However, there are some limitations to consider. Self-report measures can be inaccurate, and future work utilizing objective data with mobile and wearable technologies^52^ will enable an unbiased, real-time and unobtrusive measurement of movement behaviours with high temporal resolution. For example, as only intense levels of physical activity are those typically linked to favorable health outcomes,^53^ the use of accelerometers/mobile technology could help quantify physical activity objectively to examine the effect of duration, type and intensity of activity on various health indicators. Secondly, we did not have information on daytime activities during the rest of the 24h day that could additionally impact on outcomes observed, such as whether certain groups engaged more in educational activities that would benefit cognition and GMV development. Third, adoption of healthy amounts of 24h-MBs in certain groups of children could reflect greater parental awareness and supervision. Parents who closely regulate these behaviours in their children are also more likely to seek out enriching activities, provide proper nutrition and spend more time with their children that would additionally impact overall health and wellbeing. Finally, due to COVID-19 restrictions beginning March 2020, some participants completed remote assessments using a variety of devices for T2 (monitored by researchers using Zoom’s screen sharing feature when possible), which could have influenced the nature of some tasks. The pandemic itself also worsened adherence to 24h movement recommendations^54,55^ due to restrictions in outdoor movement and could have independently affected these health indicators.^56^ Nonetheless, separate analyses conducted on the subset of children who completed T2 assessments before March 2020 yielded similar conclusions; for data completeness we chose not to exclude any subjects in the final analyses.

## CONCLUSIONS

In conclusion, children aged 9-11y who met 24h movement recommendations of both sleep and screen time consistently demonstrated better cognition, psychosocial health, lower levels of adiposity and greater gray matter volumes than those who did not meet these recommendations. These associations persisted in longitudinal analyses conducted 2 years later suggesting the importance of maintaining these behaviours for overall health and wellbeing. Positive associations between moderate amounts of weekly physical activity and health outcome measures also suggest a need for further refinements into its optimal amount.

## Data Availability

The ABCD data used in this report came from the Curated Annual Release 4.0, available upon request from the NIMH Data Archive (https://nda.nih.gov/abcd).

http://dx.doi.org/10.15154/1523041

## Abbreviations

24h-MB: 24h movement behaviour
BMI: body mass index
GMV: gray matter volume
ICV: intracranial volume
MVPA: moderate-to-vigorous physical activity

## ACKNOWLEDGEMENTS

Data used in the preparation of this article were obtained from the Adolescent Brain Cognitive Development (ABCD) Study (https://abcdstudy.org), held in the NIMH Data Archive (NDA). This is a multisite, longitudinal study designed to recruit more than 10,000 children age 9-10 and follow them over 10 years into early adulthood. The ABCD Study®□ is supported by the National Institutes of Health and additional federal partners under award numbers U01DA041048, U01DA050989, U01DA051016, U01DA041022, U01DA051018, U01DA051037, U01DA050987, U01DA041174, U01DA041106, U01DA041117, U01DA041028, U01DA041134, U01DA050988, U01DA051039, U01DA041156, U01DA041025, U01DA041120, U01DA051038, U01DA041148, U01DA041093, U01DA041089, U24DA041123, U24DA041147. A full list of supporters is available at https://abcdstudy.org/federal-partners.html. A listing of participating sites and a complete listing of the study investigators can be found at https://abcdstudy.org/consortium_members/. ABCD consortium investigators designed and implemented the study and/or provided data but did not necessarily participate in the analysis or writing of this report. This manuscript reflects the views of the authors and may not reflect the opinions or views of the NIH or ABCD consortium investigators.

## DATA STATEMENT

The ABCD data repository grows and changes over time. The ABCD data used in this report came from the Curated Annual Release 4.0, also defined in NDA Study 1223 (doi:10.15154/1522595). All de-identified participant data are available upon request from the NIMH Data Archive (https://nda.nih.gov/abcd).

