## Supplemental Material for "Adherence to 24h movement recommendations and health indicators in the ABCD study: Cross-sectional and longitudinal associations with cognition, psychosocial health, BMI and brain structure"

**Supplementary Table 1.** The number of children included for each domain after excluding missing data and structural brain data that did not pass quality control checks.

| **Description** | **T1** | **T2** | **T2-T1** |
| --- | --- | --- | --- |
| Released data | 11,876 | 11,876 | 11,876 |
| Complete info for age (T1/T2), sex (T1), race (T1), highest parent education (T1), household income (T1) | 10,597 | 9,361 | 9,361 |
| Complete info for 24h movement behaviours | 10,574 | 9,273 | 9,255 |
| Complete info for fluid intelligence | 10,377 | - | - |
| Complete info for crystallised intelligence | 10,422 | 6,733 | 6,636 |
| Complete info for mean cognitive score (5 tasks) | 10,455 | 9,009 | 8,895 |
| Complete info for psychosocial health | 10,571 | 7,275 | 7,262 |
| Complete info for BMI data | 10,482 | 6,709 | 6,664 |
| Complete info for structural brain MRI | 10,479 | 7,046 | 6,991 |
| Passed T1w image quality checks | 10,160 | 6,930 | 6,684 |

**List of Supplementary Figures**

**Supplementary Figure 1** Post-hoc analyses showing associations between adherence to 24h movement recommendations and lobar GMVs (frontal, parietal, temporal, occipital). Plots show unstandardized estimates of regression coefficients from cross-sectional and longitudinal multilevel models for baseline (T1; left panel) and follow-up (T2; right panel) respectively. Error bars indicate 95% confidence intervals while asterisks denote level of significance (‘***’ p< 0.001 ‘**’ p < 0.01 ‘*’ p< 0.05). The gray reference line refers to the group who did not meet any recommendations. All models were controlled for age, sex, highest level of education (parent), race, household income and intracranial volume. Families nested within site were additionally modelled as a random effect

**Supplementary Figure 2** Top panel: Histogram of participants who met sleep and screen time guidelines at baseline (T1) and follow-up (T2). Bottom panel: Longitudinal (T2-T1) associations between adherence to sleep and screen time recommendations and (A) cognitive scores, (B) psychosocial health scores, (C) BMI and (D) gray matter volumes (GMV). The gray reference line refers to the group who did not meet sleep and screen time recommendations at both time points. Error bars indicate 95% confidence intervals while asterisks denote level of significance (‘***’ p< 0.001 ‘**’ p < 0.01 ‘*’ p< 0.05). All models were controlled for age, sex, highest level of education (parent), race, household income, intracranial volume (GMV models only) as well as time elapsed between baseline and follow-up and baseline measures of all dependent variables. Families nested within site were additionally modelled as a random effect.

**Supplementary Figure 3** Post-hoc analyses showing associations between frequency of physical activity and cognition (left panels), cortical GMV (middle panels) and externalizing behaviours (right panels) for T1 (top panels) and T2 (bottom panels) respectively. Plots show unstandardized estimates of regression coefficients from cross-sectional multilevel models with 0 days/week as the reference level. Error bars indicate 95% confidence intervals while asterisks denote level of significance (‘***’ p< 0.001 ‘**’ p < 0.01 ‘*’ p< 0.05). All models were controlled for age, sex, highest level of education (parent), race, household income, whether or not sleep recommendations were met (0/1 indicator variable), whether or not screen time recommendations were met (0/1 indicator variable) and intracranial volume (GMV models only). Families nested within site were additionally modelled as a random effect.
