## Supplementary figures and images for "Adherence to 24h movement recommendations and health indicators in the ABCD study: Cross-sectional and longitudinal associations with cognition, psychosocial health, BMI and brain structure"

### Figure S1

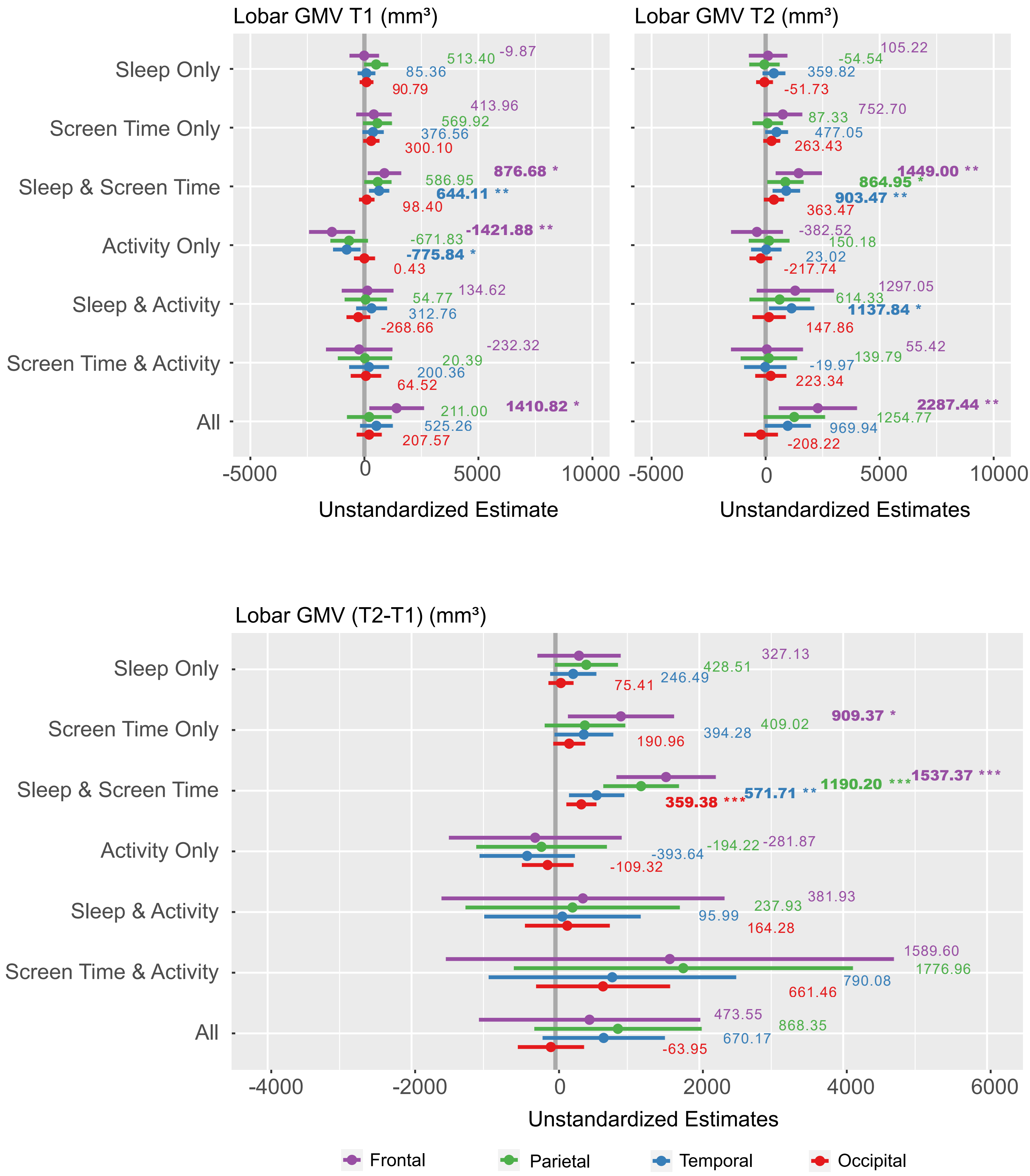

### Figure S2

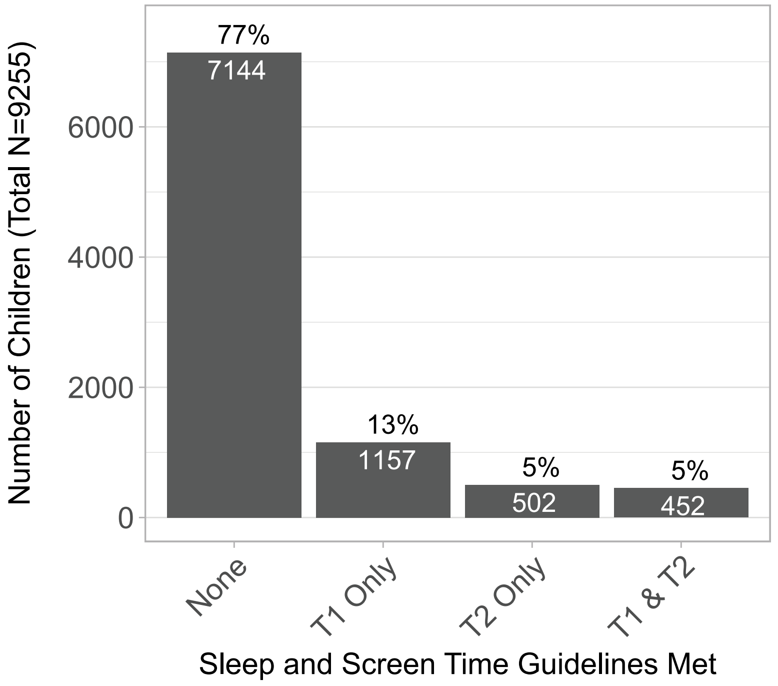

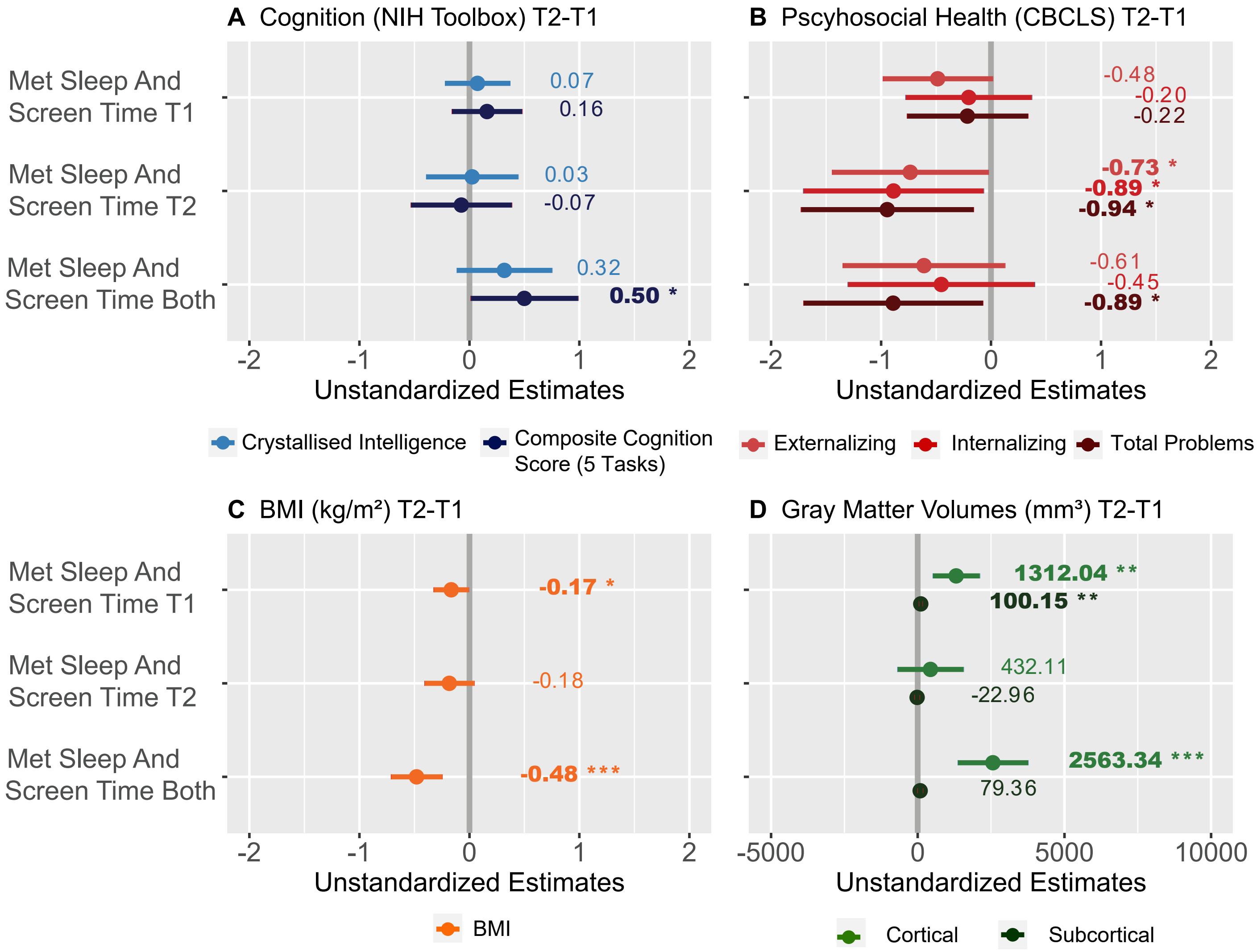

### Figure S3

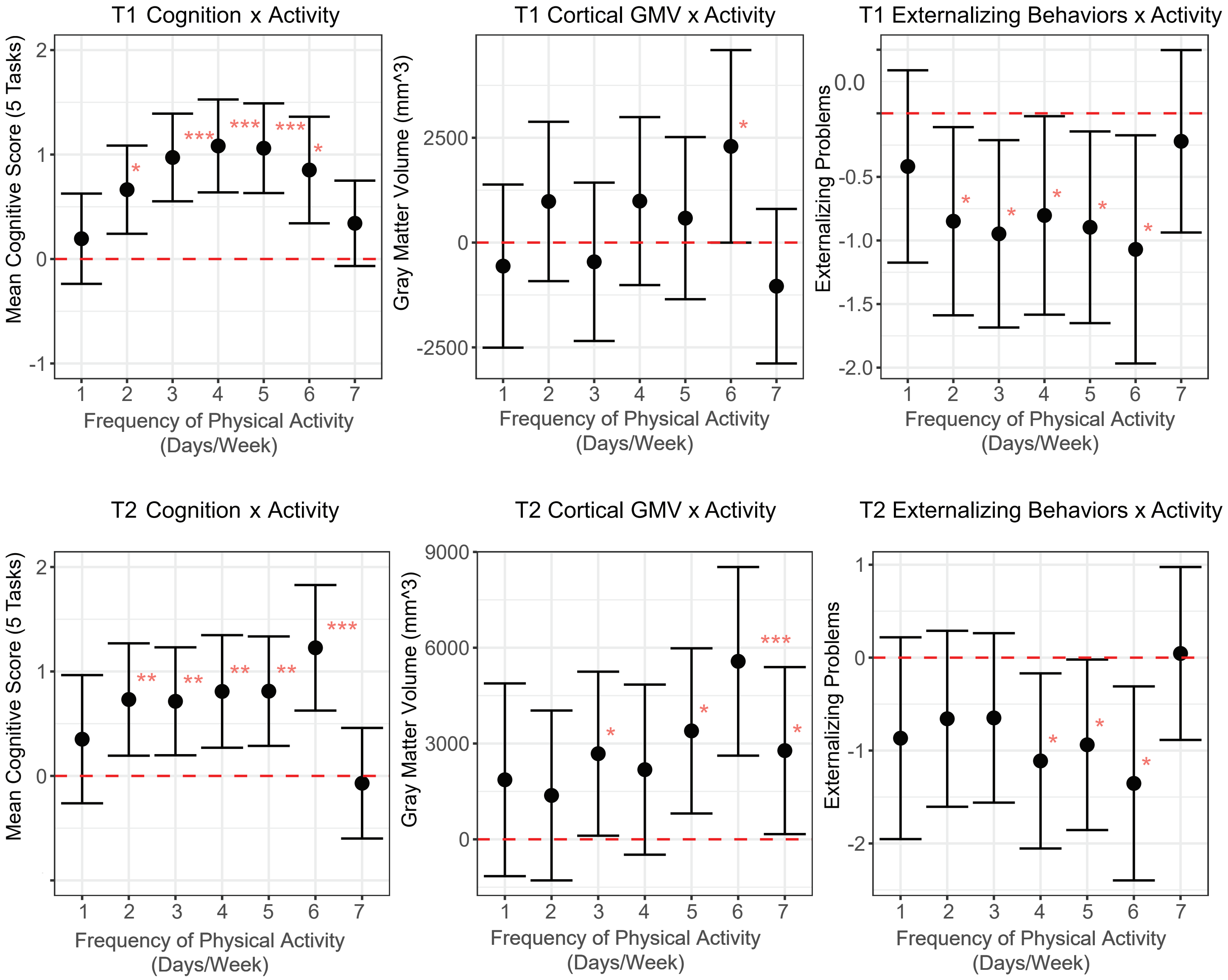
